# The Spectre of SARS-CoV-2 in the Ambient Urban Natural Water in Ahmedabad and Guwahati: A Tale of Two Cities

**DOI:** 10.1101/2021.06.12.21258829

**Authors:** Manish Kumar, Payal Mazumder, Jyoti Prakash Deka, Vaibhav Srivastava, Chandan Mahanta, Ritusmita Goswami, Shilangi Gupta, Madhvi Joshi, AL. Ramanathan

## Abstract

COVID-19 positive patients can egest live SARS-CoV-2 virus and viral genome fragments through faecal matter and urine, raising concerns about viral transmission through faecal-oral route and/or contaminated aerosolized water. These worries are heightened in many low and middle income nations, where raw sewage is often dumped into surface waterways and open defecation betide. In this manuscript, we attempt to discern the presence of SARS-CoV-2 genetic material (ORF-1ab, N and S genes) in two urban cities of India viz., Ahmedabad, in western India with several WWTPs; and Guwahati in the north-eastern part of the country with no such treatment plants. The study was carried out to establish applicability of WBE for COVID-19 surveillance as a potential tool for public health monitoring at the community level. 25.8% and 20% of the surface water samples had detectable SARS-CoV-2 RNA load in Ahmedabad and Guwahati, respectively. The high concentration of gene (ORF-1ab – 800 copies/L for Sabarmati river, Ahmedabad and S-gene – 565 copies/L for Bharalu urban river, Guwahati) found in natural waters indicates WWTPs do not always completely remove the genetic material of the virus. The study shows the applicability of WBE surveillance of COVID-19 in cities with low sanitation as well as in rural areas. The method used in this study cannot detect the live viruses, hence further research is required to evaluate the transmission implication of COVID-19 via ambient water, if any.

**Graphical abstract:** 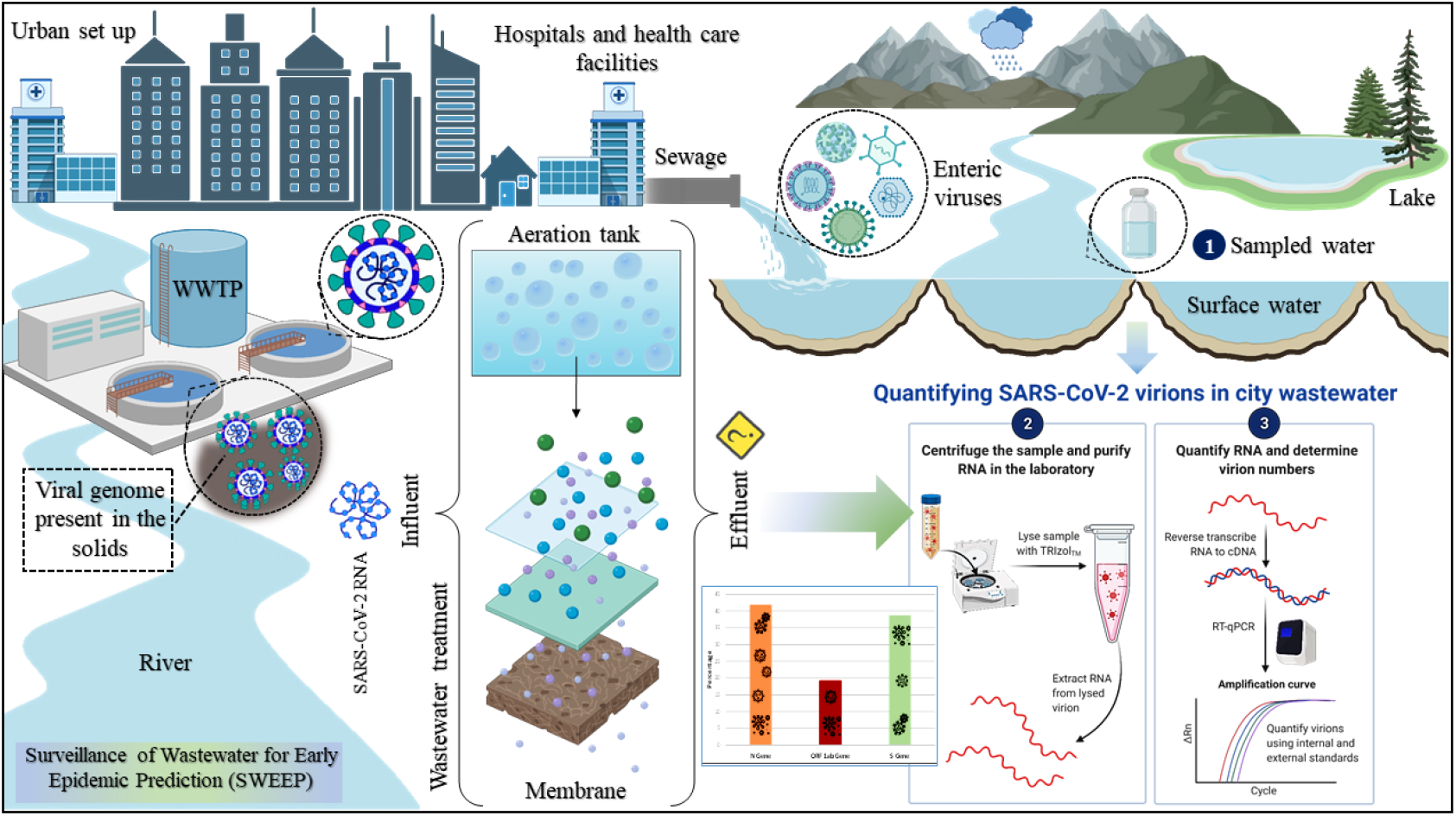

**Highlights:**

- Natural urban waters show the presence of SARS-CoV-2 RNA.
- Lake water receiving runoff containing SARS-CoV-2 genes reflected positive sign early
- Viral RNA in surface water reflects incomplete removal of gene fragments in WWTPs.
- Residence time and fate owing to viral RNA in natural waters needs further research.

## Introduction

Viruses are reported to occur in the surface water and believed to impact environmental and human health (**Lu and Yu, 2018; Qu et al., 2018, Kauppinen et al., 2018; Sekwadi et al., 2018; Kuroda et al., 2015; Kumar et al., 2019; Kumar et al., 2020**). Absence of sufficient sewage collection and treatment system is likely to make the situation more severe, especially in cities of the developing countries owing to high population density, discharge of (often unregulated) domestic and industrial effluents and ineffective treatment of wastewater (**Samaraweera et al., 2019**). It is a known fact that enteric viruses can enter into the aquatic environments through several routes such as water outflows or heavy rainfall, combined sewer outflows, blockages or sanitation system failures (**Fong et al., 2010; Kumar et al., 2019**). The coronavirus that causes severe acute respiratory syndrome (SARS) can infect the gastrointestinal system and shed in the environment, allowing for human-to-human transmission **(Ding and Liang, 2020)**. SARS are also reported to be prevalent in wastewater and surface water despite being an enveloped virus, that rapidly degrade in the environment.

The prevalence of such viruses in the aquatic environment is likely to increase considerably during the ongoing Coronavirus disease (COVID-19) pandemic situation, that pose severe health risk to humans via faecal-oral transmission or aerosolisation of water droplets containing virus (**Lodder and de Roda Husman, 2020; Naddeo and Liu, 2020**), also knowing the viable viral particles might be particularly important for Quantitative Microbiological Risk Assessment (QMRA) associated to exposure to SARS-CoV-2 contaminated water. Nonetheless, because numerous countries like India have witnessed the largest second wave of COVID-19 peaks and a probable forthcoming future wave, it has become highly imperative to adopt the wastewater based epidemiology (WBE) for early prediction of the infection. Overall, considering the millions of infections and deaths related to COVID-19, it is highly pertinent to monitor the occurrences of SARS-CoV-2 in the freshwater and wastewater systems which is vital for human sustenance. However, faecal shedding of the virus and its detection in wastewater might be particularly problematic in low-sanitation areas where wastewater treatment is partial or non-existent **(Kozer et al., 2021; Guerrero-Latorre et al., 2020)**.

Further, the abundance of viruses in tropical countries has not been well documented. As, the lipids of viral envelop can be easily disrupted by environmental stressors (**Pinon and Vialette, 2018**), enveloped viruses such as SARS-CoV-2 are more susceptible than non-enveloped viruses (e.g. Norovirus, Rhinovirus, etc.) under similar adverse environmental/system conditions (**Gundy et al., 2009**). Although, the high temperatures and solar radiations during tropical summers can effectively lower the prevalence of viruses, COVID-19 spread in the world does not suggest such (**Carratala et al., 2013; Baker et al., 2021**). The pathway of SARS-CoV-2 genes reaching to the ambient waters have been plenty (**Kumar et al., 2020**), including that of short circuiting of wastewater release into the urban waters and incomplete removal of viruses during treatment. It was found that tertiary treatment of wastewater could remove greater % of SARS-CoV-2 RNA (100%) than that of secondary treatment (89%) (**Randazzo et al., 2020**). **de Oliveira et al, (2021)** detected SARS-CoV-2 in artificially spiked river water (filtered and unfiltered) at two different temperatures viz., 4°C and 24°C through plaque assays.

On the other hand, **Haramoto et al, (2020)** reported no positive results for virus RNA in raw wastewater whereas, ∼2400 gene copies/L were detected in wastewater with secondary treatment. They also sampled surface water (river) to detect the viral genome, however, there was no trace of SARS-CoV-2 RNA in river water. Surprisingly, they also observed the abundance of N genes in positive secondary treated samples but ORF-1a and S genes were not found. Although the frequency of reports on SARS-CoV-2 presence in the treated wastewater is increasing day by day (**Westhaus et al., 2021, Hasan et al., 2021**), the ambient urban waters are somehow not being monitored. Hence it is very likely that we are going to miss this opportunity to learn a lot about the pandemic situation to make our future generations capable of understanding and manage them better. **Mancuso et al, (2021)** reviewed how SARS-CoV-2 might infiltrate the urban water cycle and subsequently spread from urban to rural water settings, posing a possible risk to crop production and, hence, human health. **Mahlknecht et al, (2021)** reported the first study on the detection of SARS-CoV-2 RNA in groundwater in Monterrey. Under the preview of the reported studies (**de Oliveira et al., 2021; Guerrero-Latorre, et. al., 2020; La Rosa, et. al., 2020; Mahlknecht, et. al., 2021; Mancuso, et. al., 2021**) from the other countries, it was vital to include surface water bodies for enhancing the prediction by WBE. There is currently no indication that COVID-19 may be transferred to animals or humans through polluted water (**La Rosa et al., 2020**). Despite this, the World Health Organization (**WHO, 2020**) has emphasised the need of study into the novel coronavirus persistence in environmental matrices like as surface water and wastewater.

Under the light of above discussion, we conducted monitoring titre of SARS-CoV-2 genetic material in various surface waters of two Indian cities i.e. Ahmedabad in Gujarat Province and Guwahati in Assam. Cities are selected such that the former has one of the highest number of wastewater treatment plants (WWTPs) among the Indian cities i.e. Ahmedabad and the latter do not have even a single treatment plant available in the city i.e., case of Guwahati. Our main objectives were to: i) understand the frequency of positive occurrence of SARS-CoV-2 RNA titre during weekly surveillance of the representative water bodies present in both the cites; ii) comparative assessment of the vulnerability of urban waters in a city setup among the silhouette of COVID-19 clinical cases. The study was carried out with the intention to check the capability of wastewater surveillance in cities with low sanitation facilities with no wastewater treatment plant(s). Can WBE approach be implemented in such cities and even in rural areas for early warning (up to 2 weeks) of COVID-19 spike in the community by sampling the ambient water? Our research is critical since there are several developing and underdeveloped nations facing poorly managed sewage systems, which result in wastewater leakages and common sewage overflow issues.

## 2. Materials and methods

### 2.1 Study area and sampling location

In the present study, three lakes i.e. Kankaria Lake, Chandola Lake, Vastrapur Lake and the Sabarmati Rivers were sampled weekly since September 3^rd^, 2020 to 29^th^ December, 2020, as a representative urban ambient water bodies in Ahmedabad **(Fig. 1a)**. In Ahmedabad, the sewage is collected through a system comprising an underground drainage network, auxiliary pumping stations (APS), Sewage Treatment Plants (STPs), and are disposed into the natural water bodies and rivers after treatment. Wastewater generated from all these development is collected by a network of underground sewers and pumping stations and is conveyed to the sewage treatment works for physical and biological treatment to meet the Gujarat Pollution Control Board (GPCB) guidelines before discharge into the nearest water body. The Ahmedabad Municipal Corporation comprises of 9 STPs, 45 Sewage Pumping Stations, and an extended Sewage Network of ∼2500 km present in the city.

**Fig.1.**
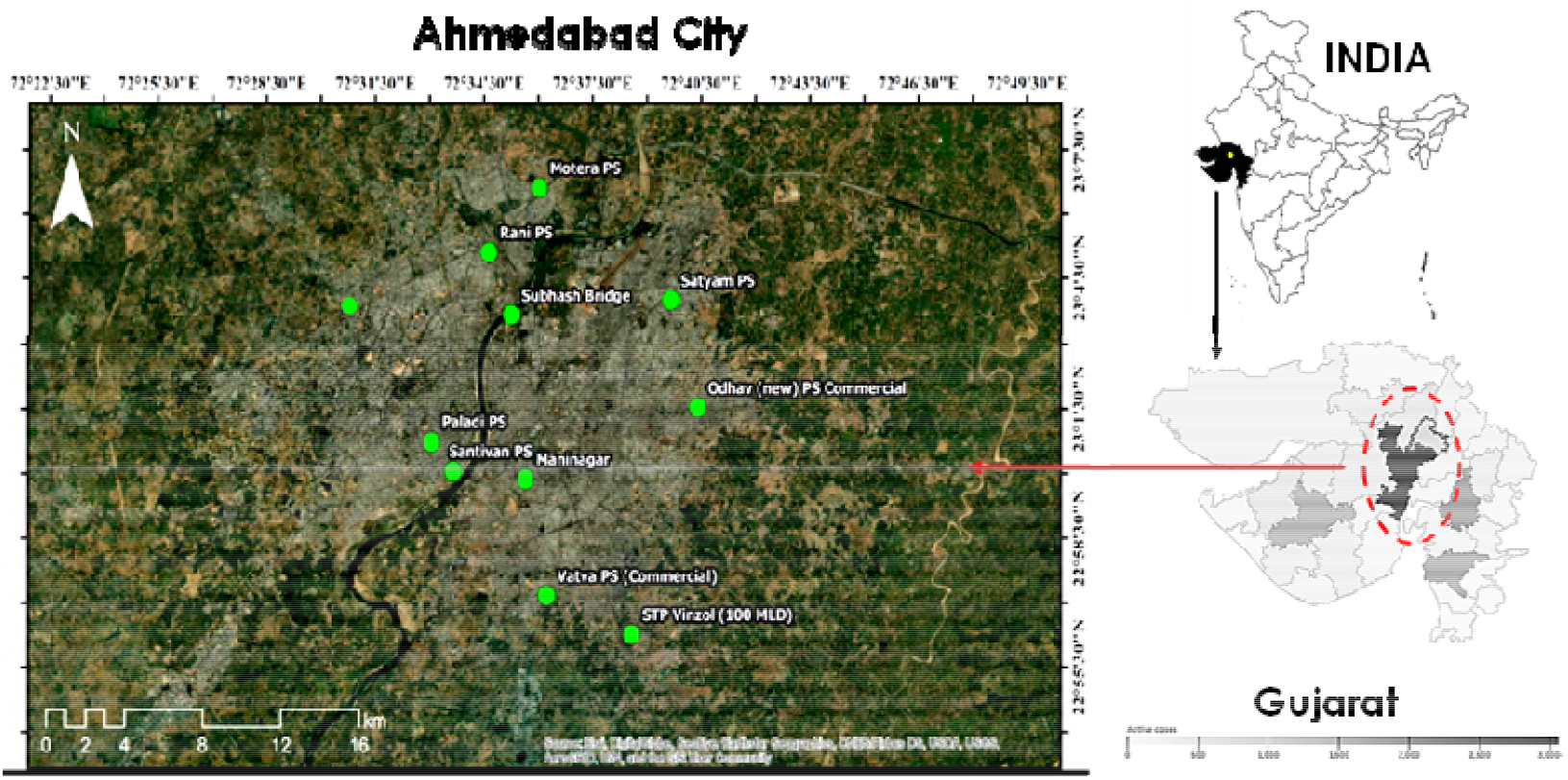
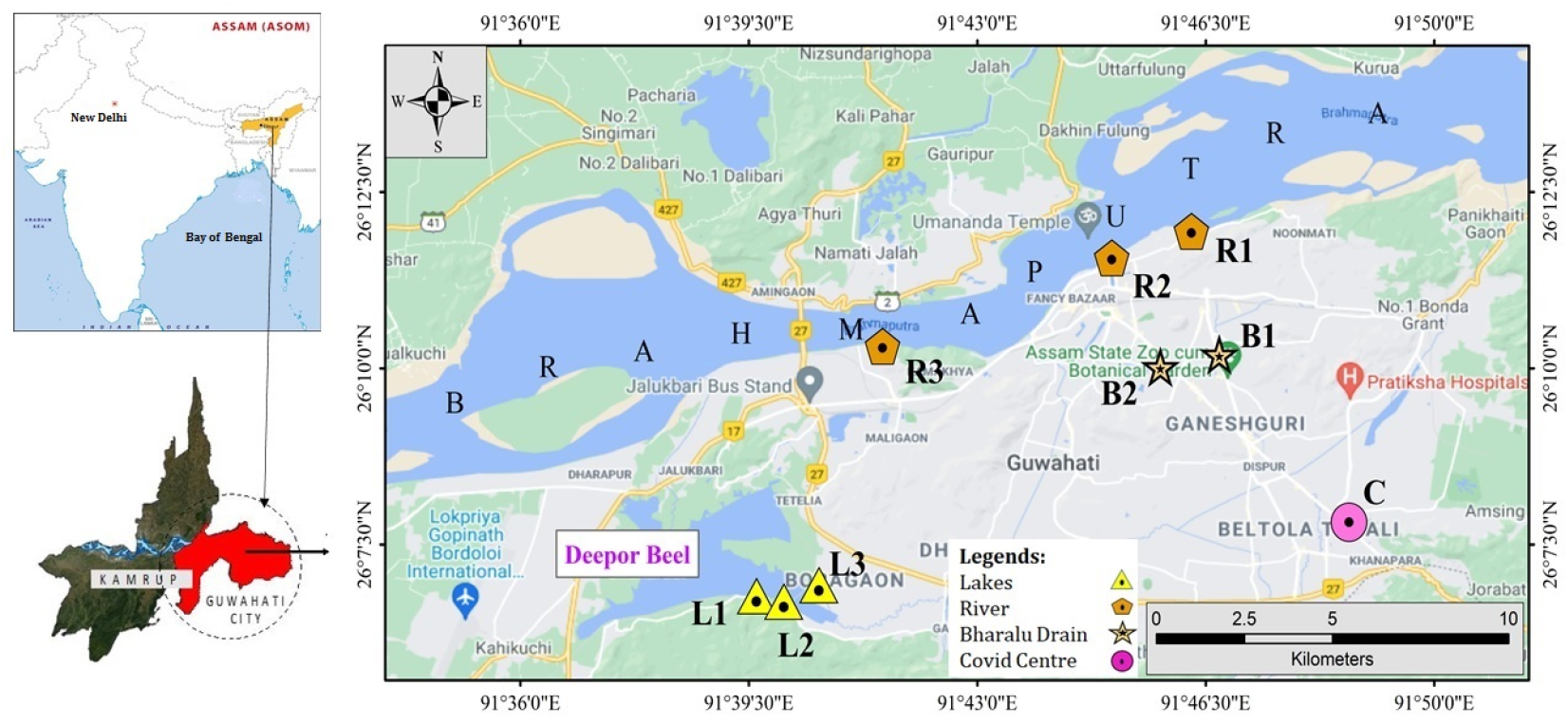
Map depicting the sampling sites in (a) Ahmedabad, Gujarat and (b) Guwahati, Assam

On the other hand, ten samples representing Dipor Bil Lake, the Brahmaputra River, the Bharalu River and the Urban Drains of the Guwahati city were taken and analysed monthly from October to December, 2020 **(Fig. 1b)**. Guwahati, known as gateway of the north-eastern India, has a concise area of 328 km^2^ that exhibit rapid and unplanned urban growth with around a million of city residents as per the 2011 census. The Brahmaputra River, an international transboundary, the fifteenth longest and the ninth largest river in terms of discharge (**Pervez and Henebry, 2015**) provides one side boundary to the city. While the Bharalu River, a tributary of Brahmaputra River, flows through the dense urban region of Guwahati city and now has virtually become an urban drain. Dipor Bil Lake is a natural freshwater lake/wetland system recognised under the Ramsar Convention provides another side of the city. There is not a single STP present in Guwahati city of Assam Province.

Probably the main solution of the wastewater here is the dilution owing to relatively higher rainfall (average annual precipitation of 2054 mm) with 91.9 average rainy days over a year. The perennial discharge of the Brahmaputra River is disposing the responsibility of diluting all the wastes of the city. Sampling locations in Guwahati was selected based on our previous work (**Kumar et al., 2019**). We added two additional locations i.e. Khanapara and AIDC based on COVID-19 quarantine centre locations in the city. Overall, eight sampling locations were precisely same as described in **Kumar et al, (2019)** and two additional locations were added specific to COVID-19 pandemic. Samples were collected using composite grab sampling by mixing three samples simultaneously taken at each location.

### 2.2 Sample collection and preparation

The samples were collected using grab sampling technique in 500ml polyethylene sterile bottles (Tarsons, PP Autoclavable, Wide Mouth Bottle, Cat No. 582240, India) and transferred in an icebox to the laboratory at Gujarat Biotechnology Research Centre (GBRC) and refrigerated at 4^0^C until further process. To take the cross-contamination during transportation into account, the sampling blanks were prepared and analysed. Samples from Guwahati was transported in a sealed ice-box by air-mail within the same day of sampling and RNA extraction was performed within 72 hrs of sampling.

Poly ethylene glycol (PEG) based precipitation method was used for concentration of the sample as explained by (**Kumar et al., 2020b**). Briefly, 30ml sample was centrifuged (Model: Sorvall ST 40R, Thermo Scientific) at 4000g for 30 minutes in a 50ml falcon tube followed by the filtration of the supernatant with a syringe filter of 0.2µ (Mixed cellulose esters syringe filter, Himedia). The filtrate was then treated with NaCl (17.5 g/L) and PEG 9000 (80 g/L) and incubated at 100 rpm overnight (Model: Incu-Shaker−; 10LR, Benchmark). The room temperature was maintained at 17 ^0^C using air-conditioner. A protocol for the same was established before and the effect of several variables like volume of the samples, temperature, rpm speed, and amount of PEG and NaCl were already observed and standardized. To make the pellet, the solution was then subjected to ultra-centrifugation at 14000g for 90 minutes (Model: Incu-Shaker™ 10LR, Benchmark). RNase-free water was used for the resuspension of the pellet containing viral particles, which then was stored in a 1.5ml Eppendorf tube at a temperature of -40°C until RNA isolation. The detailed work flow concept has been depicted in **Fig. 2**.

**Fig.2.**
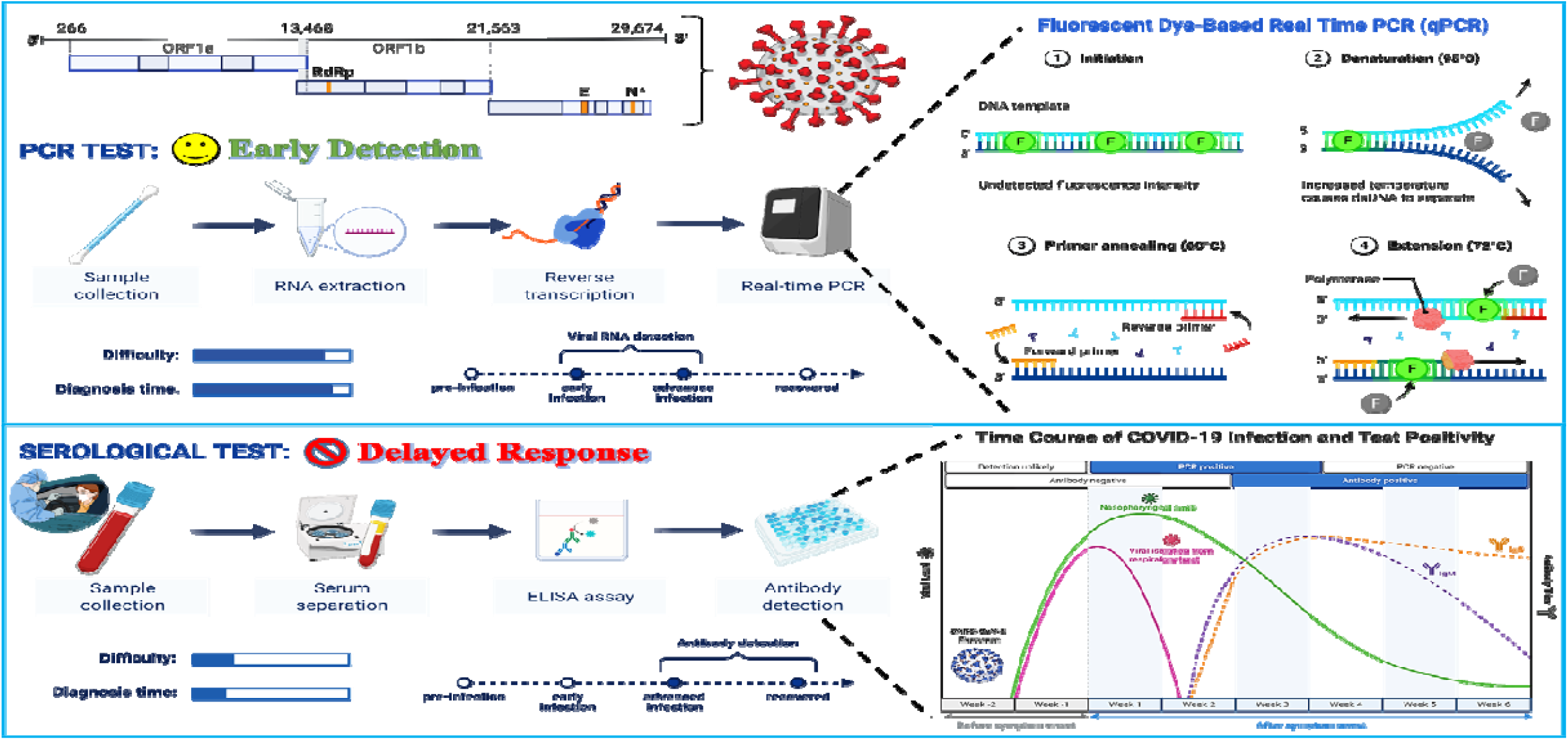
Advantage of qRT-PCR based detection of SARS-CoV-2 RNA over clinical tests for early detection, prediction and management of COVID-19 pandemic

### 2.3 Isolation of the SARS-CoV-2 viral genome

SARS-CoV-2 RNA isolation was performed using a commercially ready-for-use kit (NucleoSpin® RNA Virus, Macherey-Nagel GmbH & Co. KG, Germany). MS2 phage (10 μL), Proteinase K (20 μL) and RAV1 buffer (600 μL) consisting of carrier RNA were mixed with 300 μL of the concentrated viral particles. MS2 phage serves as the molecular process inhibition as a test control. It was used to monitor the efficacy of RNA extraction and PCR inhibition. It should be remembered that MS2 may spontaneously exist in wastewater, so there is a risk that the retrieved MS2 may consist of both the spiked and the background viral material. As per the user manual instructions (Macherey-Nagel GmbH & Co. KG), further procedures were carried out. The last elution was done with 30 μL of kit-supplied elution buffer. Using a Qubit 4 Fluorometer (Invitrogen), RNA concentrations were checked.

The nucleic acid was analyzed to identify the S gene, N gene, and ORF1ab of SARS-CoV-2 and the internal control (MS2) with the help of RT-PCR using the TaqPath™ Covid-19 RT-PCR package (Applied Biosystems). Amplification was conducted in a reaction (25 μL) vial containing 7 μL of RNAs derived from each sample. 2 μL of the positive control (TaqPath™ COVID-19 Control) and refined 5 μL of negative control were used for the study. Nuclease-free water was applied as a template-free control in this analysis. Additional process steps were executed, as defined in the product guidebook. The RT-qPCR step consisting of 40 cycles, included UNG incubation (25 °C for 2 min), reverse transcription (53 °C for 10 min), and activation (95 °C for 2 min). The reactions were conducted and elucidated as instructed in the handbook of Applied BiosystemsTM 7500 Fast Real-Time PCR.

### 2.4 Data visualization

OriginPro 2019b software has been used for data analysis and to draw boxplots.

## 3. Results and discussion

Wastewater samples collected from surface urban waters of Ahmedabad (Sabarmati River, Kankaria, Chandola and Vastrapur lakes), Gujarat, India, revealed a considerable variation in SARS-CoV-2 genome titre. Analogy of qRT-PCR assay analysis for the determination of the virus genetic material (N, S, and ORF 1ab genes) showed 25.8% (8/31) positive samples. The average N-gene copies were found to be maximum in Sabarmati River (694 copies/L), followed by Kankaria (549 copies/ L) and Chandola (402 copies/L) while, Vastrapur did not show the presence of N-gene. The ORF 1ab-gene copies were found maximum in samples collected from Sabarmati River (800 copies/ L), followed by Kankaria (87 copies/L). Chandola and Vastrapur lake samples were negative for the ORF-1ab gene. Similarly, the S-gene copies climbed down from: Sabarmati River (490 copies/L)> Vastrapur (58 copies/ L)> Chandola (52 copies/ L)> Kankaria (45 copies/L). Correspondingly, a higher SARS-CoV-2 genome concentration was observed in Sabarmati River (492 copies/L), followed by Kankaria (318 copies/ L) and Chandola lake sample (75 copies/L) **(Table 1a)**. The number of active COVID-19 cases in Ahmedabad on the day of sampling matched the gene amplification and detection patterns (viral genetic load) in surface water rather well **(Fig. 3)**. The N-gene was detected in many samples even though the samples were negative for ORF-1ab gene and S-gene. This may be due to the fact that there may be sparse concentration of RNA for gene specific amplification. The box plots for Ahmedabad shows highest detection of N-gene, S-gene, ORF-1ab gene and genome concentrations in copies/L for the month of November 2020 and April 2021 **(Fig. 4)**. It is also found that lake system being a closed system would not provide meaningful information and would not enhance WBE capabilities. It is why our study continued the Sabarmati River monitoring but stopped the lake water monitoring after a month.

**Table 1a:**
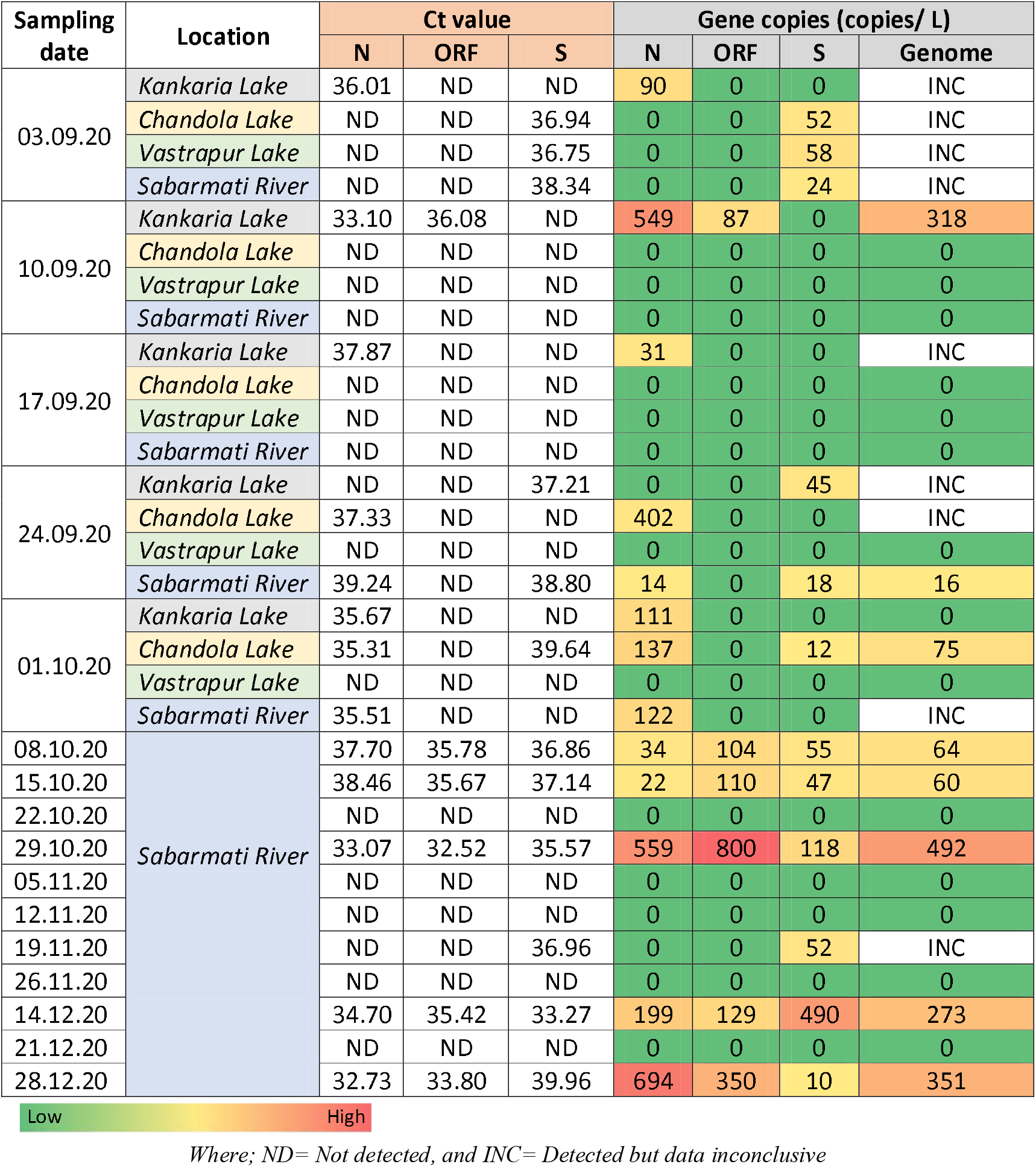
Occurrence of SARS-CoV-2 RNA traces in the freshwater samples collected from different locations in Ahmedabad.

| Sampling date | Location | Ct value |  |  | Gene copies (copies/ L) |  |  |  |
| --- | --- | --- | --- | --- | --- | --- | --- | --- |
|  |  | N | ORF | S | N | ORF | S | Genome |
| 03.09.20 | Kankaria Lake | 36.01 | ND | ND | 90 | 0 | 0 | INC |
|  | Chandola Lake | ND | ND | 36.94 | 0 | 0 | 52 | INC |
|  | Vastrapur Lake | ND | ND | 36.75 | 0 | 0 | 58 | INC |
|  | Sabarmati River | ND | ND | 38.34 | 0 | 0 | 24 | INC |
| 10.09.20 | Kankaria Lake | 33.10 | 36.08 | ND | 549 | 87 | 0 | 318 |
|  | Chandola Lake | ND | ND | ND | 0 | 0 | 0 | 0 |
|  | Vastrapur Lake | ND | ND | ND | 0 | 0 | 0 | 0 |
|  | Sabarmati River | ND | ND | ND | 0 | 0 | 0 | 0 |
| 17.09.20 | Kankaria Lake | 37.87 | ND | ND | 31 | 0 | 0 | INC |
|  | Chandola Lake | ND | ND | ND | 0 | 0 | 0 | 0 |
|  | Vastrapur Lake | ND | ND | ND | 0 | 0 | 0 | 0 |
|  | Sabarmati River | ND | ND | ND | 0 | 0 | 0 | 0 |
| 24.09.20 | Kankaria Lake | ND | ND | 37.21 | 0 | 0 | 45 | INC |
|  | Chandola Lake | 37.33 | ND | ND | 402 | 0 | 0 | INC |
|  | Vastrapur Lake | ND | ND | ND | 0 | 0 | 0 | 0 |
|  | Sabarmati River | 39.24 | ND | 38.80 | 14 | 0 | 18 | 16 |
| 01.10.20 | Kankaria Lake | 35.67 | ND | ND | 111 | 0 | 0 | 0 |
|  | Chandola Lake | 35.31 | ND | 39.64 | 137 | 0 | 12 | 75 |
|  | Vastrapur Lake | ND | ND | ND | 0 | 0 | 0 | 0 |
|  | Sabarmati River | 35.51 | ND | ND | 122 | 0 | 0 | INC |
| 08.10.20 | Sabarmati River | 37.70 | 35.78 | 36.86 | 34 | 104 | 55 | 64 |
| 15.10.20 |  | 38.46 | 35.67 | 37.14 | 22 | 110 | 47 | 60 |
| 22.10.20 |  | ND | ND | ND | 0 | 0 | 0 | 0 |
| 29.10.20 |  | 33.07 | 32.52 | 35.57 | 559 | 800 | 118 | 492 |
| 05.11.20 |  | ND | ND | ND | 0 | 0 | 0 | 0 |
| 12.11.20 |  | ND | ND | ND | 0 | 0 | 0 | 0 |
| 19.11.20 |  | ND | ND | 36.96 | 0 | 0 | 52 | INC |
| 26.11.20 |  | ND | ND | ND | 0 | 0 | 0 | 0 |
| 14.12.20 |  | 34.70 | 35.42 | 33.27 | 199 | 129 | 490 | 273 |
| 21.12.20 |  | ND | ND | ND | 0 | 0 | 0 | 0 |
| 28.12.20 |  | 32.73 | 33.80 | 39.96 | 694 | 350 | 10 | 351 |

Low High
Where; ND= Not detected, and INC= Detected but data inconclusive

**Fig.3.**
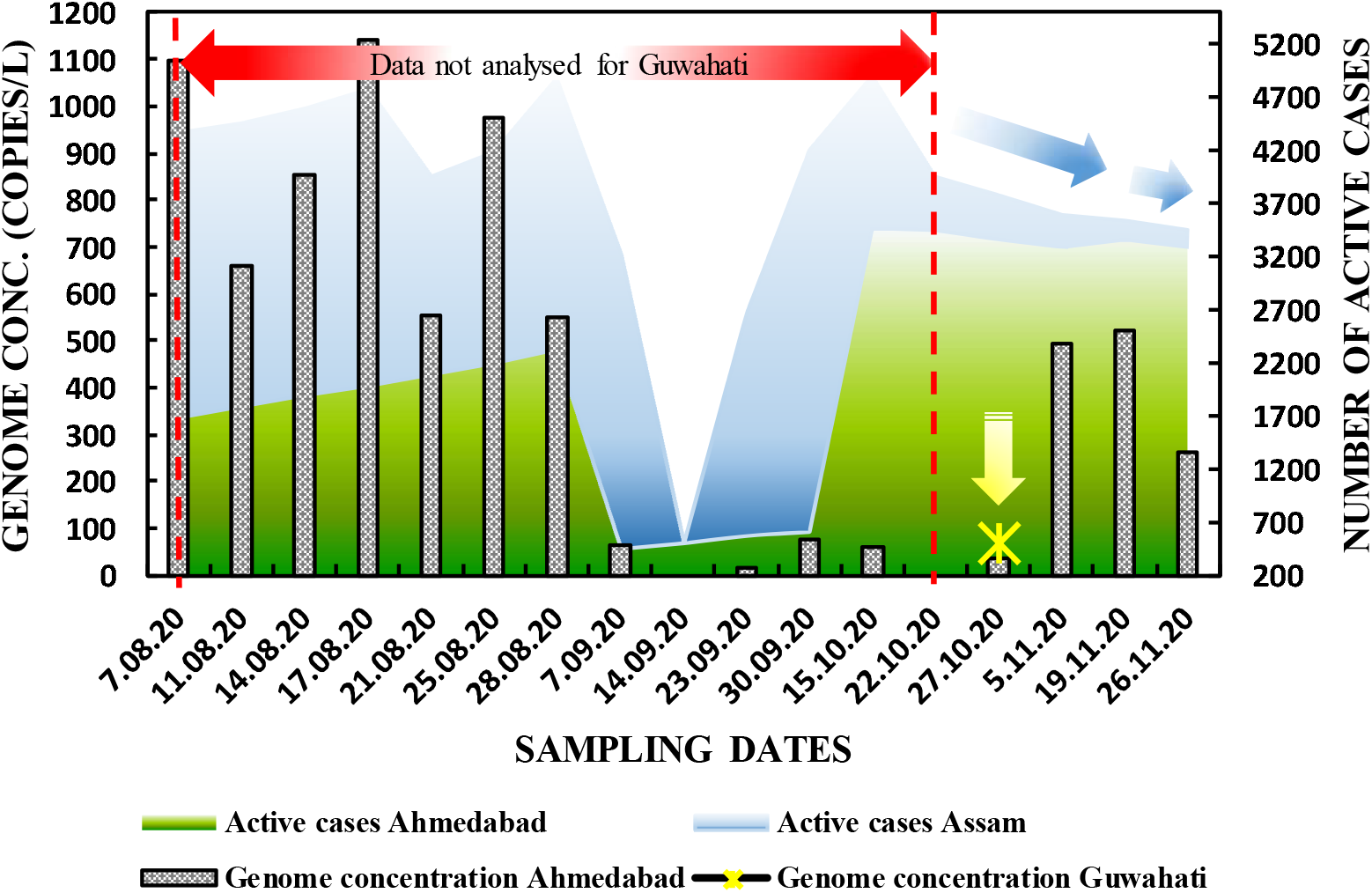
SARS-CoV-2 genome concentration as compared to clinical positive active cases in Ahmedabad and Guwahati, India

**Fig. 4.**
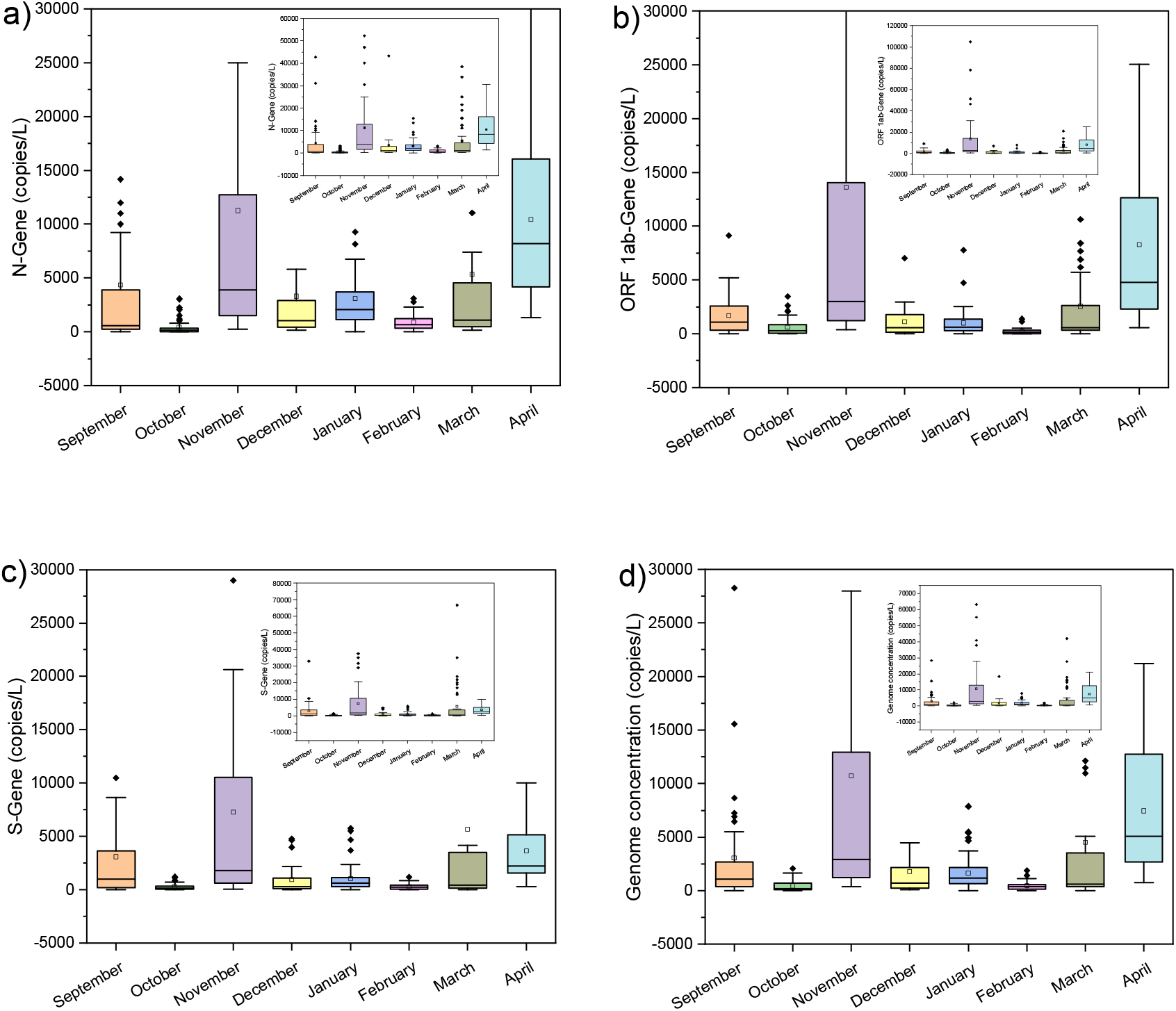
Box and whiskers plots of SARS-CoV-2 (a) N gene, (b) ORF 1ab gene, (c) S gene, and (d) genome concentration in Ahmedabad, Gujarat

The water samples collected from Guwahati (Dipor Bil lake, Brahmaputra river and WWTP at Indian Institute of Technology Guwahati (IITG) showed negative results for SARS-CoV-2 RNA. While, 1 sample near a COVID care centre and 1 sample from Bharalu urban drain tested positive for the presence of the virus genome thus showing 20% (2/10) positive results for the sampled locations. The average N-gene, ORF-1ab gene and S-gene copies were found to be maximum in the COVID care centre i.e., 9169, 4153 and 3580 copies/L than that of Bharalu urban river. However, in the Bharalu drain the S-gene concentration was found to be the highest (565 copies/L) followed by N-gene (549 copies/L) and ORF-1ab gene (435 copies/L). Evidently, a larger genome concentration was observed in the COVID care facility (5634 copies/L) than the urban drain (516 copies/L) **(Table 1b)**. Conversely, the number of active COVID-19 cases rapidly decreased in the month of October, 2020 in Guwahati which, followed the trend till March, 2021 before another rise in cases from April, 2021. The reason for negative detection of the SARS-CoV-2 gene in the Guwahati samples correspond to the decrease in clinical cases during the sampling period which, seems to be one of the lowest in the year 2020-May, 2021.

**Table 1b:** SARS-CoV-2 gene concentration in wastewater samples collected from Guwahati.

| Sampling date | Location | Ct Value |  |  | Gene copies (copies/ L) |  |  |  |
| --- | --- | --- | --- | --- | --- | --- | --- | --- |
|  |  | N-Gene | ORF-Gene | S-Gene | N-Gene | ORF-Gene | S-Gene | Genome concentration |
| 27.10.20 | Dipor Bil (Boragaon) | ND | ND | ND | ND | ND | ND | ND |
|  | Khanapara | 29 | 30.1 | 30.3 | 9169 | 4153 | 3580 | 5634 |
|  | AIDC | 33.1 | 33.5 | 33.1 | 549 | 435 | 565 | 516 |
|  | Uzan Bazar | ND | ND | ND | ND | ND | ND | ND |
|  | Dipor Bil-1 | ND | ND | ND | ND | ND | ND | ND |
|  | Bhangaghar | ND | ND | ND | ND | ND | ND | ND |
|  | Kharguli | ND | ND | ND | ND | ND | ND | ND |
|  | Pandu | ND | ND | ND | ND | ND | ND | ND |
|  | Dipor Bil-2 | ND | ND | ND | ND | ND | ND | ND |
|  | WW/IITG | ND | ND | ND | ND | ND | ND | ND |

High Low

The COVID care centre (CCC) showed positive results as the symptomatic and asymptomatic patients were treated there. Bharalu drain, however, flows through the heart and lungs of the city and collects sewage and waste before finally joining to the Brahmaputra River. Hence, the asymptomatic cases or those who were not admitted to CCC still shedding the viral RNA would be detected in the wastewater. The Bharalu river which turned into an urban drain carrying viral genetic material can play as a hub and core of surveillance for future pandemic like situations. The current results reveal the microbiological implications of sewage discharge into natural streams without prior treatment. Guwahati’s urban waterways shows viral RNA signatures possibly due to the unmediated exude of sewage water from a population of about one million people. WWTPs can remove SARS-CoV-2 RNA, thus, strengthening sanitation and health infrastructure. As per the published fact that wastewater treatment plant does not remove SARS-CoV-2 genes and/or gene fragments completely, the study adds another dimension to the wastewater surveillance and recommends monitoring of river waters.

The world is on the verge of facing a third wave of COVID-19 and India is facingmany natural calamities in 2021 e.g., several severe earthquakes in Assam and cyclone Tauktae near Gujarat coast. In such case access to safe water, health and hygiene during rehabilitation is pivotal. Therefore, all possible exposure pathways of SARS-CoV-2 RNA is needed to be considered scientifically and point of discharge needs to identified and tested for microbial contamination along with basic water quality parameters. The findings of our study implies that WBE may be applied to other cities and even rural areas as well where, sewage is disposed directly into natural waterways. It is crucial to note, however, that in the current study, only SARS-CoV-2 genetic material has been identified in waterways, and the virus’s survival in contaminated waterways is unknown. Furthermore, because zoonotic spill over episodes are common in the Coronaviridae family, viral propagation into the environment has an undisclosed influence on domestic animals and wildlife health (**Franklin & Bevins, 2020**). Eventually, if diagnostic equipment’s are restricted, the abundance of the viral genome can be employed as a surveillance criteria for a prompt warning system monitoring main sewage discharges across the city, assisting in the containment of the pandemic (**Bivins et al., 2020**).

## 4. Conclusion

The persistence of the SARS-CoV-2 virus and the viral RNA in various water matrixes is a current research subject. In the context of intermittent lockdown and progressive rise in COVID cases in India, we attempted to investigate the occurrence of SARS-CoV-2 genetic signature in two metropolitan cities of India viz., Ahmedabad (Western zone) and Guwahati (North-Eastern zone). The sustenance of the viral RNA load in urban surface waters in both the cities were congruent to the trends in active clinical COVID-19 cases. Lack of complete removal of viral RNA via wastewater treatment might be a contributing reason to its detection and elevated probability of surveillance of COVID-19 via monitoring ambient waters. Water safety begins with the preservation of natural water resources in the watershed; as a result, it is crucial to keep surface and groundwater from contamination with faeces and to prevent direct discharge of grey water into rivers, streams, lakes, wetlands, open wells, etc. Surface waters receiving direct sewage or effluent discharge can be targeted for surveillance of SARS-CoV-2 genome and thus, can provide a lot of insights on myths in transmissions, need of sanitation, probable future risks and efficient management. The approach described in this paper can be employed in other places where sampling sewage is impossible and wastewaters or treated effluents are disposed into lakes, streams or rivers. The knowledge is also helpful to indicate thorough investigation of possibility of contagion in places with inadequate sanitation, where people are at risk of being exposed to polluted water or even raw sewage. The present study is likely to contribute in the advancement of pandemic surveillance science and to support WBE applicability in urban areas without wastewater treatment plants as well as in the rural areas.

## Data Availability

All data is included in the paper.

## Acknowledgement

This work is funded by UNICEF, Gujarat and UKIERI. We also acknowledge the help received from Mr. Alok Thakur, and other GBRC staffs who contributed towards sample and data analyses.

